# Maternal morbidity profile in hospitalizations in the Unified Health System in São Paulo, Brazil: analysis using data mining, 2014 to 2019

**DOI:** 10.1101/2025.04.02.25325149

**Authors:** Rejane Sobrino Pinheiro, Marcos Augusto Bastos Dias, Luís Guilherme Buteri Alves, Luís Carlos Guillén, Lana Meijinhos, Natália Santana Paiva, Valeria Saraceni, Claudia Medina Coeli, Rosa Maria Soares Madeira Domingues

**Author notes:** **Corresponding author:** (RSP).

## Abstract

In Brazil, approximately 80% of births are registered in the Hospital Information System of the Unified Health System (SIH/SUS), which has several fields for recording morbidities. SIH/SUS is the only Brazilian information system that has morbidity data, but its use for maternal morbidity surveillance is still limited. The objective of this study was to identify a set of diagnoses that alone or together increase the risk of maternal death, via data mining technique. Obstetric hospitalizations at SIH/SUS of women aged 10-49 years from the largest state in Brazil (São Paulo) from 2014 to 2019 were analysed. Diagnoses were classified into 12 groups, adapting the World Health Organization’s classification proposal of maternal deaths. Groups of diagnoses that were related to maternal death were identified using association rules with the Apriori algorithm. Of the 2,742,467 hospitalizations, 831 (0.03%) resulted in death. The most frequent diagnoses associated with death, alone or in combination, were nonobstetric complications (62.0%). Pregnancy-specific hypertensive complications (20.1%), pregnancy-related infections (19.9%) and haemorrhages (13.6%) were also present in hospitalizations resulting in death. The risk of death was more significant for nonobstetric complications, unknown causes and external causes, with lifts above 10. The risk of death was at least three times greater for groups of diagnoses related to the most frequent causes of maternal death in the country (complications of hypertension, infections, and haemorrhages). Uncomplicated abortion, gestational diabetes and other causes of nonobstetric hospitalization were only associated with maternal death when together with other diagnoses. The results of this study reaffirm the importance of hypertensive, haemorrhagic and infectious causes for maternal death and highlight the relevance of nonobstetric causes for the surveillance of maternal morbidity since women with these diagnoses, alone or associated with other complications, yield to a higher risk of death.

## Introduction

Brazil has a high maternal mortality ratio, which is greater than 50 per 100,000 live births. In the period 2009-2020, the leading causes of maternal death were direct obstetric causes, predominantly hypertension, haemorrhage, puerperal infection, and abortion. Among the indirect obstetric causes, the main ones are diseases of the circulatory system, respiratory system diseases, and maternal infectious and parasitic diseases [1].

Reducing maternal mortality depends on detecting diseases and complications developed during pregnancy, childbirth, and the postpartum period and the consequent response of the health system to manage them, covering aspects of patient safety and the ability to reduce negative consequences. Surveillance of maternal morbidity associated with the worsening of women’s conditions during pregnancy, childbirth, or the postpartum period allows for a deeper understanding of preventable maternal deaths. It contributes to detecting deficiencies in health systems to reorient and prioritize actions.

The investigation of maternal deaths was established in Brazil in 2009 and is essential for a better understanding of the causes and determinants of maternal deaths. However, maternal deaths are infrequent, especially in the context of health services, and it is recommended that maternal morbidity be studied in a complementary manner to develop strategies to improve obstetric care and reduce maternal mortality [2].

In Brazil, approximately 80% of births are publicly funded [3] and are registered in the Hospital Information System of the Unified Health System (from the Portuguese name Sistema de Informação Hospitalar do Sistema Único de Saúde - SIH/SUS). Each hospital admission record has several fields that allow the reporting of different obstetric diagnoses and related comorbidities.

Analysis strategies using data mining can identify a set of causes that can act alone or together to increase the risk of maternal death. Thus, the objective of this study was to apply data mining techniques to analyse the profile of morbidities associated with maternal death registered in the SIH/SUS.

## Method

### Data source

We analysed the obstetric hospitalizations of women aged 10-49 years admitted to São Paulo State’s public or private hospitals funded by the Unified Health System (SUS) and registered in the SIH/SUS between January 2014 and December 2019, before the start of the COVID-19 pandemic. São Paulo is the largest state in the country and has a wide, publicly funded network of health services trained in childbirth and postpartum care.

In the SIH/SUS, a hospital admission may consist of one or more claim records, called Hospital Admission Authorization (AIH-from Portuguese, *Autorização de Internação Hospitalar*). When AIH discharge was a hospital stay or an interhospital transfer, the subsequent record (within a one-day maximum difference between the admission date and the previous discharge date) was searched via a deterministic record linkage algorithm using variables available in deidentified databases: the hospital code, the municipality of residence, and the patient’s date of birth. All AIH records of the same patient identified by the algorithm were grouped and analysed as one hospital episode. The source code of the algorithm is available at GitHub: https://github.com/kencamargo/cuidados_multiplos/tree/main/processaih. We selected hospital epsiodes where at least one AIH presented: i) an obstetric diagnosis (chapter XV of the International Classification of Disease – ICD X) in the principal or any of the 11 secondary diagnostic fields; or ii) an obstetric principal care procedure. The AIH anonymized database is publicly available at https://datasus.saude.gov.br/transferencia-de-arquivos/.

We also considered only hospital episodes where the reason for discharge was “discharged alive” or “death in hospital,” as the objective was to analyse the profile of diagnoses registered in the SIH/SUS associated with maternal death. We also excluded 222 hospital episodes with “death in hospital” as the reason for discharge because we could not identify deaths in the mortality database via a deterministic matching algorithm.

The final database analysed included 2,742,467 hospital records. Most of them (72.6%) had only one primary diagnosis and only one AIH (98.9%).

### Morbidity classification

We classified each diagnosis into 12 groups, some of which were divided into subgroups (S1 Table). Our classification was based on the one proposed by the International Classification of Diseases, 10th revision—maternal ICD of the World Health Organization (WHO) [4], which classifies maternal deaths. As the AIH database contains data on morbidity, not causes of mortality, the WHO maternal ICD groups were adapted to capture all diagnoses recorded during hospitalization, not just causes of death. The diagnoses of Group 10 (childbirth) were not included, as they are not relevant to the morbidity analysis.

### Data mining

We used a data mining technique based on association rules to identify diagnosis groups associated with hospital discharge (alive or death). The associations were assessed by considering both isolated and combined diagnoses.

This technique has been used in shopping basket analysis to identify which items in the basket are purchased together [5,6]. Association rules are based on combinatorial analysis of sets of different items belonging to a database. An item represents a category of a variable. For example, male is one item, and female is another item. The rule is based on the relationship of a set of items called antecedents (represented by X) to another set of items called consequents (represented by Y). The relationship can be expressed by a conditional sentence of the type IF (X) THEN (Y), symbolized by X→Y. An association rule depicts the relationship between items, checking whether items x_1_, x_2_,…,x_n_ are associated with items y_1_,…y_k_ [5].

The rules are generated considering all possible combinations of items, and metrics related to the association’s frequency and “strength” are calculated. The most commonly used metrics are support, confidence, and lift, which are used to select the most relevant rules.

Support is related to the frequency of a database profile (set of items). The support of a rule, Supp(X→Y), is the proportion in which the antecedents and consequents are found concomitantly in the database (Eq. 1). The confidence of a rule measures the proportion in which the consequent appears in the set of records that have the profile given by the antecedents (Eq. 2). Lift measures the dependence between the items of a rule, pointing to the strength of the association observed between the consequent and the antecedent, compared with the situation in which the consequent and the antecedent would represent independent events (Eq. 3).

In the analysed database, death in hospitals was a very infrequent event, so the minimum support and confidence were set at 1.0x10^-10^. This allowed the generation of infrequent rules to be mined later via the metrics described by Eq. (1) to (3).

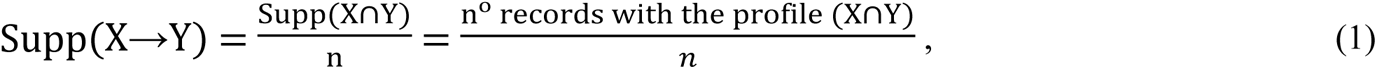

where *n* is the number of records in the database.

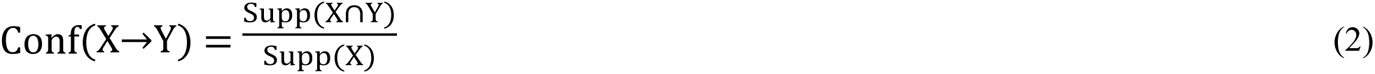

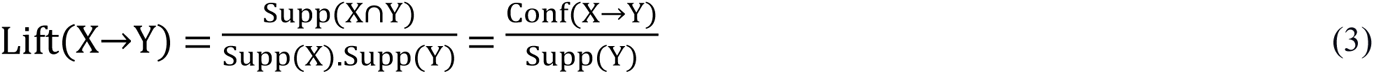

We used the Apriori algorithm [6] implemented in the arules R package (version 1.6-8) (https://www.rdocumentation.org/packages/arules/versions/1.6-8). Rules with a lift lower than 1.1 and redundant rules were eliminated via the is.redundant function and the confidence metric. A redundant rule has additional items but with confidence equal to or lower than that obtained by the original rule. The rules that contain the most relevant diagnosis groups were initially selected via support, confidence, and lift metrics. Because in obstetric hospitalization, death in the hospital is a rare event, we analysed infrequent rules when they occurred at least four times in the database. Finally, we excluded rules with diagnoses of “group 10 (childbirth)”, as they are not relevant to the morbidity analysis.

We analysed databases that were deidentified and made publicly available (open access). According to the Brazilian National Health Council ethics resolution n° 510/2016 (April 7, 2016), research ethics committee approval was waived.

## Results

Among the 2,742,467 hospital episodes evaluated, 831 (0.03%) resulted in death in the hospital as the reason for discharge.

The algorithm generated 744 rules. We eliminated all rules with discharge alive as consequent because they presented a lift less than 1.1 and analysed 120 (14.8%) cases of death in the hospital as a consequence and a lift above 1.1. Among the latter, 16 presented only one diagnosis group/subgroup in the antecedent (Table 1). The most frequent diagnosis group was nonobstetric complications (group 8), which were present in 62.0% (515) of hospital episodes that ended in death. In this group, “other nonobstetric causes” appeared in 23.7% of hospital episodes (197), respiratory complications in 15.9% (132), and circulatory complications in 14.5% (121). Pregnancy-specific hypertensive complications (group 2a) were present in 20.1% (167) of hospitalizations resulting in death, pregnancy-related infections (group 4a) in 19.9% (165), haemorrhages (group 3) in 13.6% (113), and other obstetric complications (group 5) in 9.5% (79). The other remaining groups were present in 14.3% of hospital episodes that ended in death: group 1b (abortion with complications), group 2b (chronic hypertensive complications), group “others” (other causes not classified in the previous groups), group 4b (other unrelated infections pregnancy) and group 12 (external causes) (Table 1).

**Table 1.** Frequency of diagnostic groups in hospitalizations with death in SIH/SUS. state of São Paulo, Brazil, 2015-2019.

| Group | Subgroup | Support | Confidence | Lift | Number of cases | Group total |
| --- | --- | --- | --- | --- | --- | --- |
| G1 – Abortions | 1b – With complications | 1,39E-05 | 0.001 | 3.029 | 38 | 38 |
| G2 – Hypertensive complications | 2a - Pregnancy specifics | 6,09E-05 | 0.001 | 4.376 | 167 | 201 |
|  | 2b - Chronicles | 1,24E-05 | 0.001 | 4.2177 | 34 |  |
| G3 - Hemorrhages |  | 4,12E-05 | 0.001 | 4.245 | 113 | 113 |
| G4 - Infections | 4a – Pregnancy-related | 6,02E-05 | 0.002 | 7.510 | 165 | 187 |
|  | 4b – Not related to pregnancy | 8,02E-06 | 0.002 | 6.300 | 22 |  |
| G5 - Other obstetric complications |  | 2,88E-05 | 0.002 | 5.774 | 79 | 79 |
| Group 7 - Diabetes | 7b - Non-gestational diabetes | 4,74E-06 | 0.0004 | 1.310 | 13 | 13 |
| G8 - Non-obstetric complications | 8a - Respiratory | 4,81E-05 | 0.022 | 73.018 | 132 | 515 |
|  | 8b - Digestive | 1,79E-05 | 0.015 | 51.045 | 49 |  |
|  | 8c – Circulatory | 4,41E-05 | 0.014 | 47.090 | 121 |  |
|  | 8d - Anemia | 5,83E-06 | 0.003 | 9.183 | 16 |  |
|  | 8e - Other non-obstetric causes | 7,18E-05 | 0.003 | 10.053 | 197 |  |
| G9 – Unknown causes |  | 4,74E-06 | 0.236 | 780.048 | 13 | 13 |
| G12 – External causes |  | 8,02E-06 | 0.006 | 20.190 | 22 | 22 |
| Group others |  | 1,20E-05 | 0.001 | 2.647 | 33 | 33 |

The diagnosis groups with the highest risk of death, with lifts above 10, were nonobstetric complications (group 8), unknown causes (group 9), and external causes (group 12) (Table 1). Hypertensive complications - group 2, infections - group 4, and haemorrhages – group 3, which correspond to the most frequent causes of maternal death in mortality statistics, presented lifts above 3 (Table 1).

Sixty-three rules combined two groups/subgroups, whereas 40 combined three (Table 2). Only one rule presented four diagnosis groups/subgroups in the antecedent.

**Table 2.**
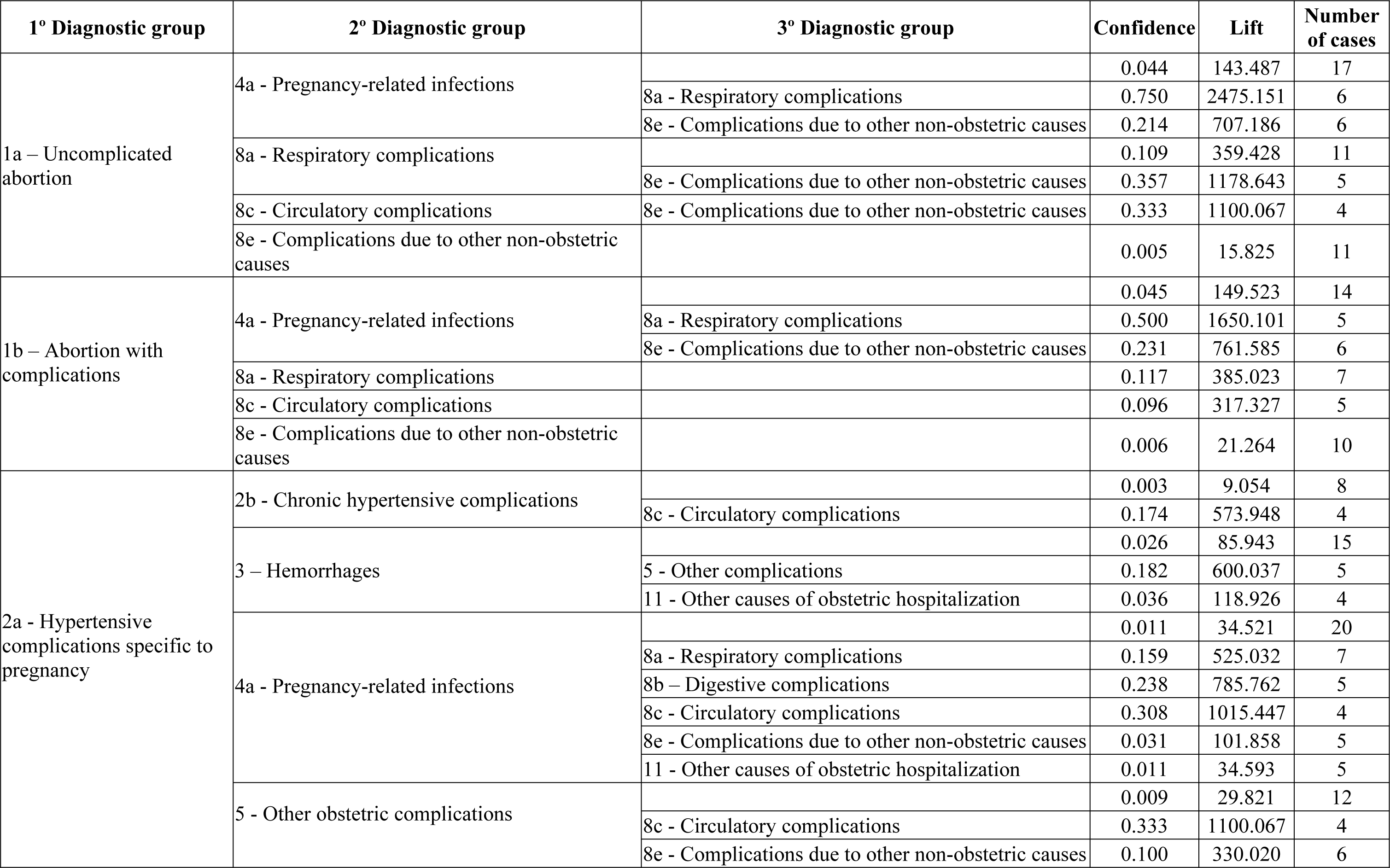
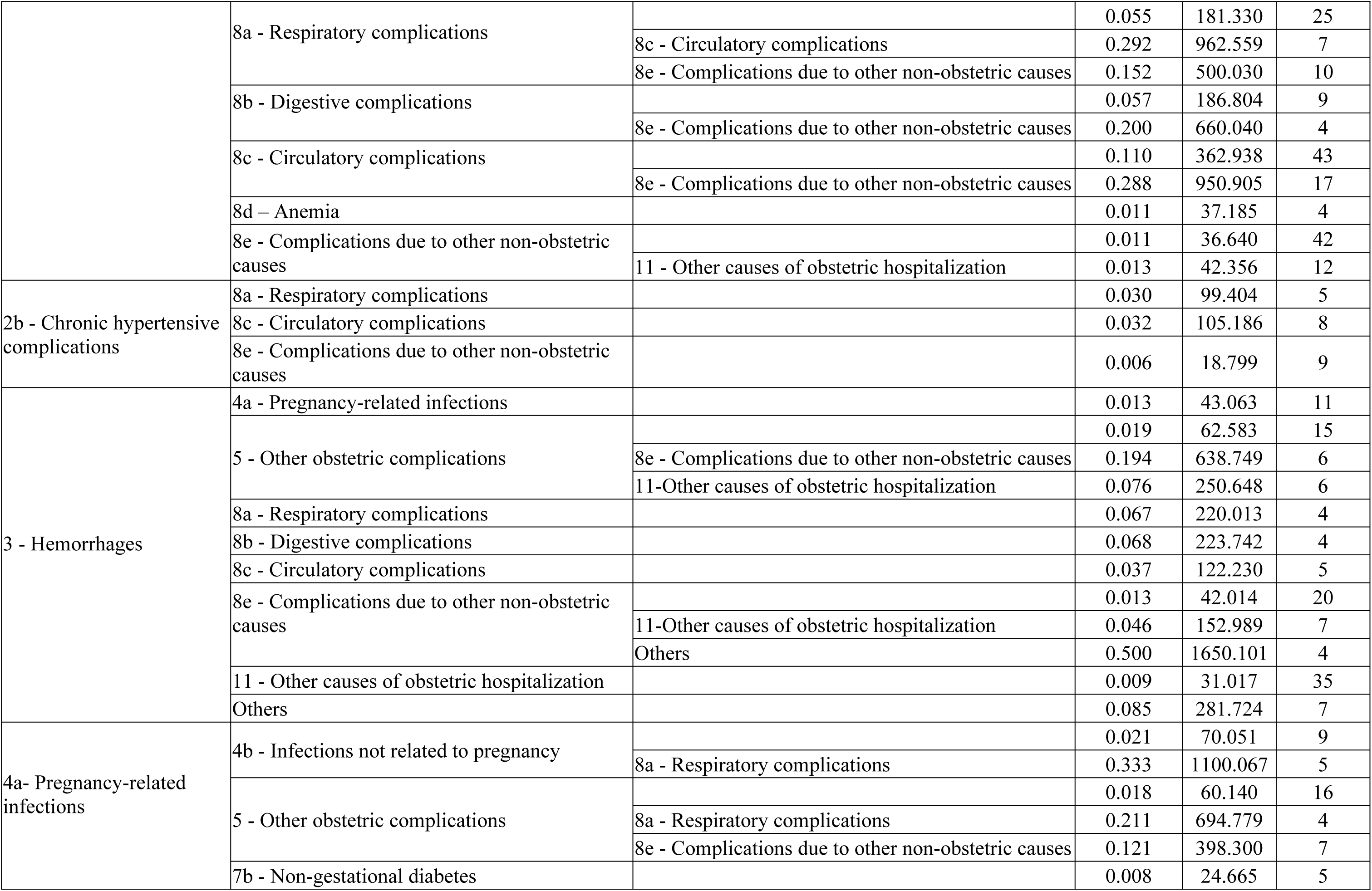
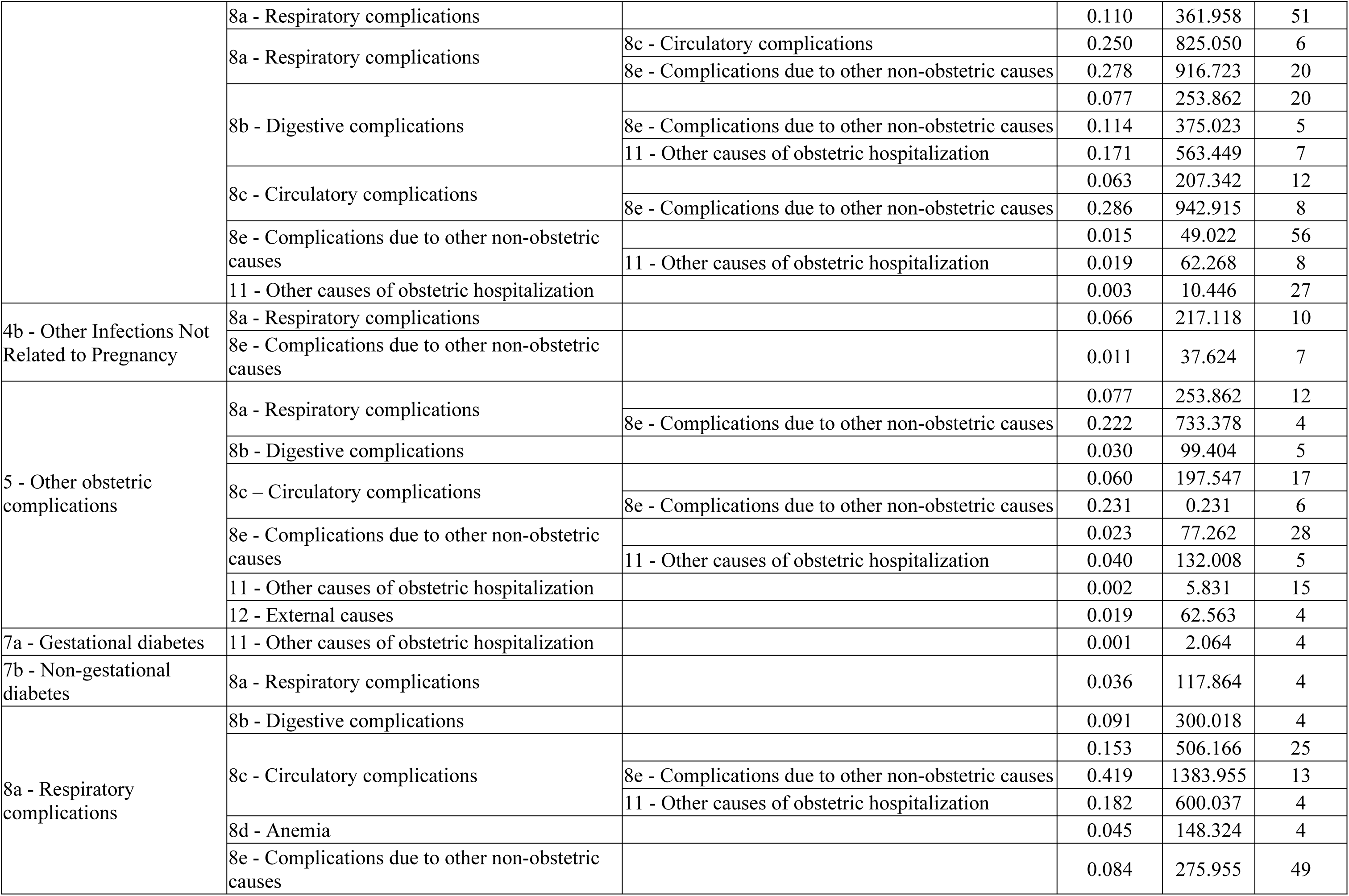
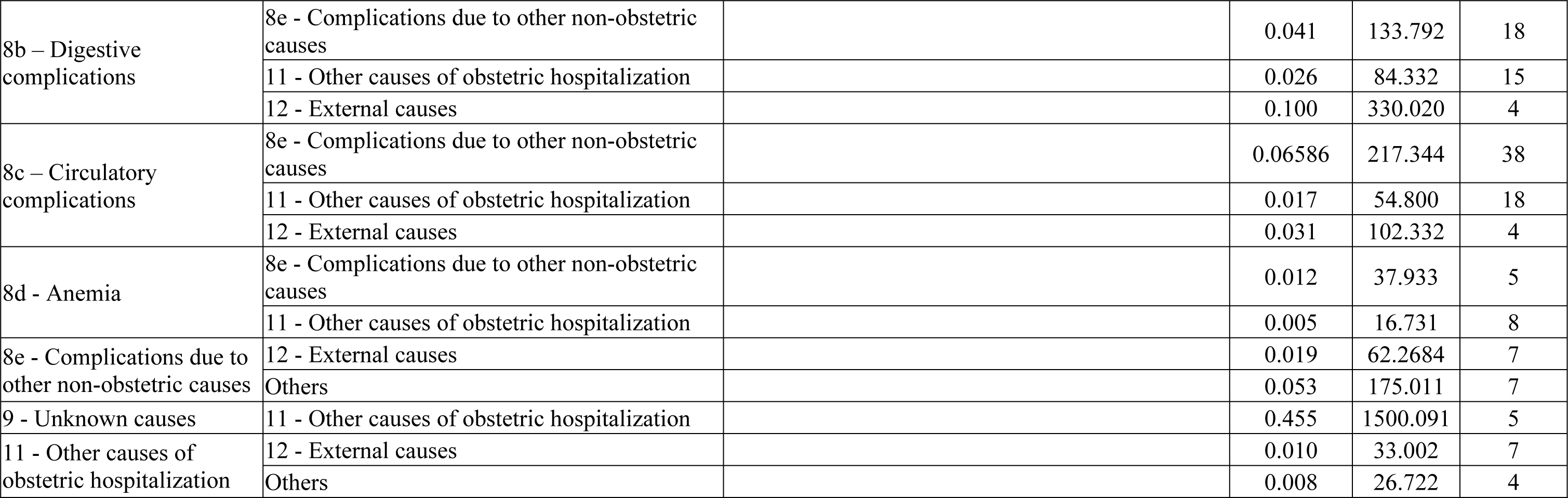
Most frequent combinations of diagnostic groups in hospitalizations that result in death. state of São Paulo, Brazil, 2015-2019.

Diagnostic groups 1a (uncomplicated abortion), 7a (gestational diabetes), and 11 (other causes of nonobstetric hospitalization) alone were not associated with maternal death. However, they appeared to be associated with death when accompanied by other diagnoses. Uncomplicated abortion (group 1a) appeared in rules together with infections related to pregnancy, respiratory or circulatory complications, or other nonobstetric causes (Table 2). Other causes of nonobstetric hospitalization (group 11) appeared together with gestational diabetes (group 7a) or other obstetric complications (group 5), external causes (group 12), and other diagnoses (group others) in hospitalizations that ended in death (Table 2).

Furthermore, Table 2 (and S1 and S2 Figs) show that when the diagnosis groups were combined with other diagnosis groups/subgroups, diagnosis groups of complications, especially the subgroup 8 (nonobstetric complications), appeared more frequently. Only groups 9 (unknown causes) and 11 (other obstetric conditions) did not occur with group 8. In general, when a subgroup was associated with group 11, the lifts were the smallest among all other combinations of that subgroup.

When a rule presents three diagnosis groups/subgroups in the antecedent, the third diagnosis group frequently includes complications from other non-obstetric causes (group 8e), increasing the risk of death by 5 to 10 times, as observed by the lift values. The third group included women with uncomplicated abortions, hypertensive complications specific to pregnancy, haemorrhages, and pregnancy-related infections (Table 2).

## Discussion

In this study, we found that nonobstetric causes, hypertensive complications, pregnancy-related infections, and haemorrhage were the diagnoses most frequently associated with death during hospitalization, whether isolated or combined. We also found that abortion and diabetes were associated with maternal death in the presence of other diagnoses, such as nonobstetric causes.

Direct obstetric diagnoses are the most common cause of death in the country. However, the group of nonobstetric causes was the most frequent group of morbidities, with very high lifts, especially for women hospitalized for abortion, nongestational diabetes and other reasons for obstetric hospitalization. This result indicates the need for greater surveillance of women with nonobstetric complications, owing to the greater risk of worsening conditions that in themselves do not pose a high risk of death and owing to the increased risk in women with other morbidities.

The interpretation of these results and their comparison with the causes of maternal death must consider the particularities of the different information systems. In the Mortality Information System, maternal deaths are classified as those occurring during pregnancy or up to the 42nd day after the end of pregnancy, due to any cause related to or aggravated by pregnancy or measures in relation to it, but not due to accidental or incidental causes. Furthermore, the definition of the cause of death occurs after the investigation of maternal death, using the cause that originated the sequence of events that resulted in death. In the SUS Hospital Information System, we consider not only deaths but also all hospitalizations where a complication was recorded, regardless of whether this complication was the main diagnosis during hospitalization or an associated cause. Furthermore, information on the date of the end of pregnancy was not available, and it is possible that deaths occurring after the 42nd day were included. Finally, external causes were also included in the morbidity analysis.

Despite these differences, we observed that the main causes of death in the country and the state of São Paulo – hypertensive causes, haemorrhage and infections – were among the most frequently recorded diagnosis groups, with lifts above three, which shows that death was three times more common in women who presented these complications. These results are consistent with the national maternal mortality profile and reinforce the need for improved surveillance of these complications.

The comparison of our results with those of other maternal morbidity studies is limited by how morbidity is measured, an aspect already highlighted by other authors [7]. In this study, only diagnoses, rather than medical procedures, were included. In addition, the analysis approach uses data mining techniques to identify the causes that alone or together increase the risk of maternal death. We did not identify studies in the literature that have adopted this same analysis strategy.

Brazilian [8,9,10] and international [11] morbidity studies that adopted the World Health Organization [12] criteria for identifying cases of maternal near miss used clinical, laboratory and management markers of organic dysfunction without identifying the underlying cause. The criteria used by the WHO for “potentially life-threatening conditions”, also called severe maternal morbidity, are based on management severity indicators and diagnoses of hemorrhagic, hypertensive and other systemic disorders [12]. Haemorrhages and hypertension are related to direct obstetric causes. Moreover, other systemic diagnoses include conditions such as sepsis, shock, pulmonary embolism, convulsions and respiratory failure, which do not allow the determination of whether the origin of the disorder was a direct, indirect or nonobstetric cause. Therefore, the contribution of nonobstetric causes becomes less evident.

The use of morbidity scores based on the existence of preexisting conditions or those diagnosed during pregnancy has been proposed to predict severe maternal morbidity [13,14]. Different conditions receive different weights depending on their association with the outcome, and higher scores show a more significant association with the outcome. One of the studies reported an increase in the proportion of pregnant women with a score greater than or equal to 1 in the USA. One of the explanations is the greater prevalence of chronic conditions in pregnant women, the presence of which, alone or in combination with other morbidities, affects pregnancy and childbirth management [13].

SIH/SUS has limitations for evaluating maternal mortality. The cause of death was present in only 13.5% of the cases and was not subject to investigation. Approximately 23.7% of deaths occurred in women whose complications were not recorded. In other words, women who were hospitalized for obstetric care with no record of morbidity but who died. This result suggests the occurrence of acute complications during hospital admission without their registration in the system. We also found very high lifts in women in group 9 (unknown causes), probably recorded as deaths with a short period of hospital stay, preventing a more precise diagnostic assessment. Data from a systematic review including 26 studies and 8,704,230 women reinforce this hypothesis by showing that 48.9% of postpartum maternal deaths occurred in the first 24 hours after birth, mainly in low- and middle-income countries. Most deaths from postpartum haemorrhage and pulmonary embolism occur in the first 24 hours postpartum (79.0% and 58.2%, respectively). On the other hand, deaths from hypertensive complications occur mainly in the first week postpartum, and deaths from infections occur between the 8th and 42nd day postpartum [15].

As positive aspects of the study, we highlight the inclusion of almost 3 million publicly funded obstetric hospitalizations that occurred in the largest state of the country; the adoption of a hospital episode approach, grouping multiple events of the same patient; the comprehensive analysis of all recorded diagnoses; and the machine learning technique used to identify associations.

## Conclusion

The study of maternal morbidity is an essential component of maternal health surveillance, contributing to improving obstetric care, as it is more frequent than maternal deaths and shares the same determinants. SIH/SUS is the only Brazilian information system with obstetric morbidity data, but its use for morbidity surveillance is still limited.

The results of this study reaffirm the importance of hypertensive, hemorrhagic and infectious causes for maternal death and highlight the relevance of nonobstetric causes for the surveillance of maternal morbidity. It is essential to identify women with these complications, which, alone or associated with other complications, are at greater risk of death.

For SIH/SUS to be used to monitor maternal morbidity, continuous improvement of the data registry is necessary, with adequate completion of all ICDs for complications occurring during hospitalization, as well as deaths and their causes. Future studies should evaluate the importance of the main causes of hospitalization and investigate the sequence of complications that result in death, contributing to a better understanding of cases and the definition of alert systems.

## Data Availability

https://datasus.saude.gov.br/transferencia-de-arquivos/

https://drive.google.com/drive/folders/1eceyf0uN6l4u-pAQpRgzSy9uvZeK3qHZ?usp=sharing

https://datasus.saude.gov.br/transferencia-de-arquivos/

## Supporting information

**S1 Table. Diagnostic groups of hospitalizations adapted using the WHO classification causes of maternal death as reference.**

**S1 Fig.**
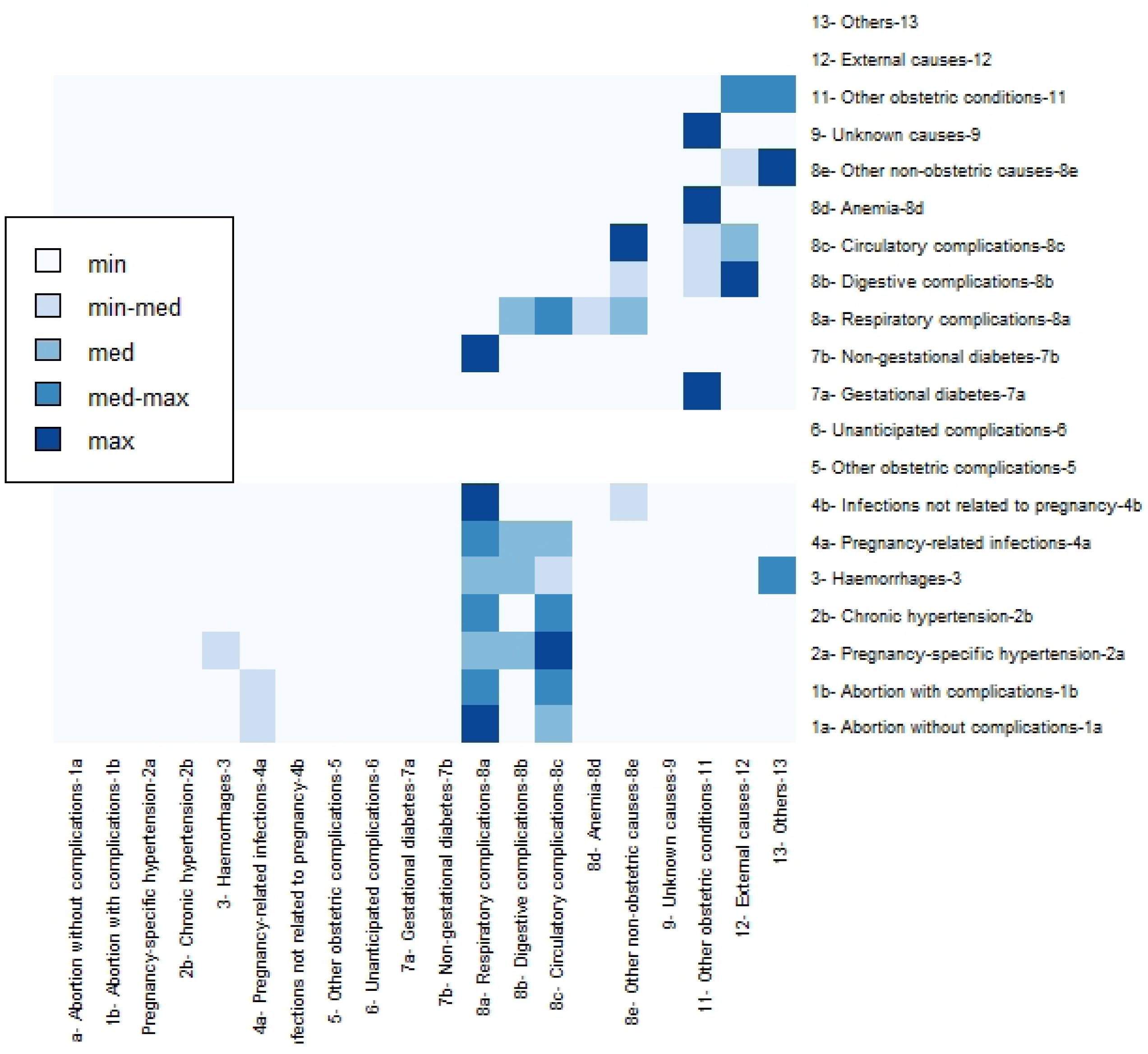
Heatmap of rule lifts related to death, with 2 diagnostic groups. Brazil, São Paulo state, 2015-2019.

**S2 Fig.**
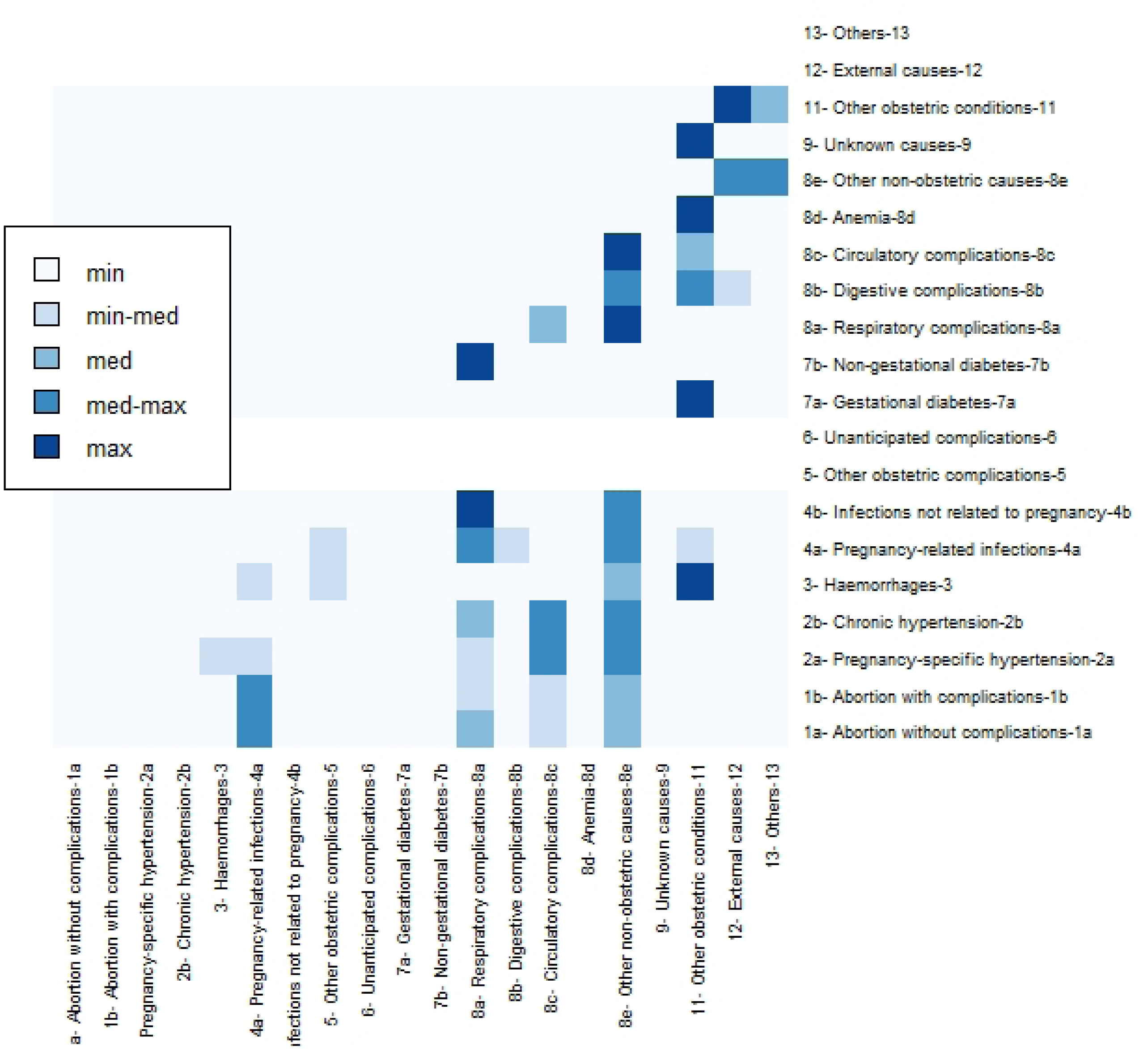
Heatmap of rule counts related to death, with 2 diagnostic groups. Brazil, São Paulo state, 2015-2019.

## References

1. Brazil. Ministry of Health, Health Surveillance Secretariat. Epidemiological Bulletin 20, vol 53, year 2022. Maternal Mortality in Brazil 2009-2020. Available at https://www.gov.br/saude/pt-br/centrais-de-conteudo/publicacoes/boletins/epidemiologicos/edicoes/2022/boletim-epidemiologico-vol-53-no20/view, accessed on 31 October 2023.

2. World Health Organization. Evaluating the quality of care for severe pregnancy complications: the WHO near-miss approach for maternal health. Geneva; 2011. https://apps.who.int/iris/bitstream/handle/10665/44692/9789241502221_eng.pdf;jsessionid=EEBF2C5BAD3A80F2E66623549925154B?sequence=1, accessed on July 11, 2023.

3. Domingues RMSM, Dias MAB, Nakamura-Pereira M, Torres JA, d’Orsi E, Pereira APE, et al. Decision process for the type of birth in Brazil: from women’s initial preference to the final route of birth. Cad Public Health. 2014;30:S101–16.

4. World Health Organization. The WHO application of ICD-10 to deaths during pregnancy, childbirth, and puerperium: ICD MM. Geneva: World Health Organization; 2012. Available at https://www.who.int/publications/i/item/9789241548458, accessed on January 2, 2024.

5. Agrawal R, XSriKant R. Fast Algorithms for Mining Association Rules. Proceedings of the 20th VLDB Conference Santiago, Chile, 1994. Access: October 18, 2023. Available at: https://www.vldb.org/conf/1994/P487.PDF.

6. Han J, Kamber M, Pei J. Data mining: concepts and techniques. 3rd. Ed. Morgan Kaufmann: Massachusetts, 2012.

7. Nam JY. Comparison of global indicators for severe maternal morbidity among South Korean women who delivered from 2003 to 2018: a population-based retrospective cohort study. Reprod Health. 2022 Aug 13;19(1):177. doi: 10.1186/s12978-022-01482-y.

8. Nakamura-Pereira M, Mendes-Silva W, Dias MAB, Reichenheim ME, Lobato G. Hospital Information System of the Unified Health System (SIH-SUS): an evaluation of its performance for identifying maternal near miss. Cad Public Health 2013; 29(7), 1333–1345.

9. Dias MA, Domingues RM, Schilithz AO, Nakamura-Pereira M, Diniz CS, Brum IR et al. Incidence of maternal near miss in hospital childbirth and postpartum: data from the Birth in Brazil study. Public Health Cad. 2014;30 Suppl 1:S1–12.

10. Cecatti JG, Costa ML, Haddad SM, Parpinelli MA, Souza JP, Sousa MH et al. Network for Surveillance of Severe Maternal Morbidity: a powerful national collaboration generating data on maternal health outcomes and care. BJOG. 2016;123(6):946–53.

11. Nelissen E, Mduma E, Broerse J, Ersdal H, Evjen-Olsen B, van Roosmalen J et al. Applicability of the WHO maternal near miss criteria in a low-resource setting. PLoS One. 2013; 8 (4): e61248. Doi: 10.1371/journal.pone.0061248.

12. Say L, Souza JP, Pattinson RC. Maternal near miss - towards a standard tool for monitoring the quality of maternal health care. Best Pract Res Clin Obstet Gynaecol. 2009; 23(3):287–296.

13. Salahuddin M, Mandell DJ, Lakey DL, Ramsey PS, Eppes CS, Davidson CM, Ortique CF, Patel DA. Maternal comorbidity index and severe maternal morbidity during delivery hospitalizations in Texas, 2011-2014. Birth. 2020;47(1):89-97. doi: 10.1111/birt.12465.

14. Leonard SA, Kennedy CJ, Carmichael SL, Lyell DJ, Main EK. An Expanded Obstetric Comorbidity Scoring System for Predicting Severe Maternal Morbidity. Obstet Gynecol. 2020 Sep;136(3):440–449. doi: 10.1097/AOG.0000000000004022.

15. Dol J, Hughes B, Bonet M, Dorey R, Dorling J, Grant A, et al. Timing of maternal mortality and severe morbidity during the postpartum period: a systematic review. JBI Evid Synth. 2022;20(9):2119–2194. doi: 10.11124/JBIES-20-00578.

